# A simplified approach to monitoring the COVID-19 epidemiologic situation using waste water analysis and its application in Russia

**DOI:** 10.1101/2020.09.21.20197244

**Authors:** Polina Alexandrovna Kuryntseva, Kamalya Oktay Karamova, Valentin Petrovich Fomin, Svetlana Yurevna Selivanovskaya, Polina Yurevna Galitskaya

## Abstract

The number of registered cases of COVID-19 is increasing in the world, and some countries are reporting a second wave of the pandemic. Accurate and real time information about epidemiological situation is therefore urgently needed for managing decisions in the countries, regions and municipalities which are affected. Massive testing of viral presence in people’s saliva, a smear from the nose, nasopharynx and / or oropharynx, bronchial lavage water obtained by fibrobronchoscopy (bronchoalveolar lavage), as well as from (endo) tracheal, nasopharyngeal aspirate, sputum, biopsy or autopsy material of the lungs, whole blood, serum or antibodies presence in blood cannot give relevant information about the COVID-19 infection rate in the community since simultaneous testing of the whole community is not technically possible, the information obtained in testing of specific groups is retarded and, in addition, such testing is expensive. The alternative to mass testing of the population is the testing of wastewater that could contain SARS-CoV-2 particles originating from excreta. Such testing has several limitations connected with the particularities of the testing procedure.

In the present study, a modified approach for detection of COVID-19 infection rate using wastewater analysis has been developed. The approach includes i) the creation of a calibration curve on the basis of the serial dilution of excreta collected from people who are infected with COVID-19 and ii) the analysis of the wastewater samples and their serial dilutions, the approach excludes usage of concentrating techniques before wastewater sample analysis as well as usage of external control in RT-PCR reactions for calculation of numbers of viral particles. The minimum infection rate that can be detected using this approach is 10-2%. The approach developed was used to investigate wastewater from eleven sewage inspection chambers in the city of Kazan (Russia). It was demonstrated that the average infection rate of people using these sewers was over 0.4% in July 2020.

## Introduction

The WHO announced that the COVID-19 outbreak had become a pandemic on 12th March 2020 WHO 2020). COVID-19 is caused by the respiratory SARS-CoV-2 virus with a long incubation period, and very different courses of disease – from severe to asymptomatic. To the 15^th^ of September 2020, over 29 million coronavirus cases have been reported in the world, and over 1 million of them in Russia, which has 4^th^ highest amount of cases in the world (https://www.worldometers.info).

Currently, the basic mechanism for determination of the COVID-19 epidemiological situation in countries is testing symptomatic cases using the PCR (polymerase chain reaction) technique and evaluating the number of positive tests over time. Additional mechanisms include analysis of symptoms such as those which are most common (fever, dry cough, tiredness) and less common symptoms (aches and pains, sore throat, diarrhea, conjunctivitis, headache, loss of taste or smell, a skin rash, or discoloration of fingers or toes) and also serious symptoms (breathing difficulty or shortness of breath, chest pain or pressure, loss of speech or movement) (“Coronavirus”; La Rosa et al., 2020; Mao et al., 2020). These techniques can be characterized as biased and displaying both resource insensitivity and detection blindness as well as having a high cost. As a result, all countries may have a delayed view of the real epidemiological picture, and less wealthy countries may receive underestimated data. Besides, the production capacity of laboratories at the peak of the epidemic cannot cope with mass testing of people in a short time. Precise and real-time information about the COVID-19 epidemiological situation is, however, extremely important for decision makers in municipalities, regions and countries.

A WBE (water-based epidemiology) approach that is based on pathogen determination in the wastewater of whole settlements or districts is a cheap and relevant alternative to this challenge. WBE assumes that the identification and quantification of pathogens in community wastewater reflects the health status of the community population in real time (WHO “Water, sanitation…”; Mao et al., 2020). It was demonstrated that SARS-CoV-2 viral particles are present in the feces of infected people, including those who are asymptomatic, in quantities up to 10^10^ copies·g^-1^ (Wu et al., 2020; La Rosa et al., 2020; Kitajima et al., 2020). Furthermore, it was shown that viral particles are present in the community’s wastewater. Thus, SARS-CoV-2 RNA was found in wastewater in the Netherlands, the USA, Australia and Italy, even before intensive growth of reported coronavirus cases numbers (Wu et al., 2020; Medema et al., 2020; Ahmed et al., 2020; La Rosa et al., 2020). The numbers of viral particles in the wastewater is reported to be 10^2^-10^4^ copies·l^-1^ (Wu et al., 2020; Wurtzer et al., 2020; Medema et al., 2020; Ahmed et al., 2020; Haramoto et al., 2020), and indirectly these numbers compared with those in the initial feces samples allow us to calculate the ratio of people in the community infected by the virus. This approach has, however, several limitations. The procedure of viral gene copy number estimation includes sample concentrating with different efficacy depending on the method and matrix (feces or wastewater), RNA extraction from different matrices containing different PCR and RT (reverse transcription) reaction inhibitors, RT and PCR reactions, both with their own efficacy depending on reagents, protocols and external controls used, and on RNA purity. Therefore, the results obtained may contain significant discrepancies. Some authors suggest relying only on qualitative (yes/no), but not on quantitative, information while analyzing COVID-19 epidemiological situation using WBE approach (Wu et al., 2020; Haramoto et al., 2020). Besides, the procedure of SARS-CoV-2 gene copy number estimation described above has limitations in terms of equipment and consumables requirements, which means that the WBE approach cannot be implemented in a daily routine immediately.

In Russia, pathogens are checked in cleaned wastewater in order to estimate the efficacy of a wastewater plant’s functioning, but not in the initial wastewater in the framework of WBE monitoring. Consequently, no information is available about the presence of SARS-CoV-2 particles in the wastewater. However, this information is of great importance not only because of WBE advantages described above but also because the number of coronavirus cases in Russia might be underestimated (“COVID-19 Situation Reports”). As well as the presence of asymptomatic cases and the shortage of testing facilities at the peak of the epidemic that are equally responsible for underestimations in other countries, it may also be connected with the local mentality. Except for severe cases, people prefer to stay home, to take several days off and not to appeal to the doctor. Together with the lifting of quarantine, this habit may cause the second wave of COVID-19 to spread; especially in the areas with high population density such as the major Russian cities: Moscow (4949 people·km^-2^), Saint-Petersburg (3847 people·km^-2^), Nizhniy Novgorod (3049 people·km^-2^), Novosibirsk (3215 people·km^-2^), Samara (2136 people·km^-2^) and Kazan (2135 people·km^-2^).

For detection of SARS-CoV-2 particles in saliva, a smear from the nose, nasopharynx and / or oropharynx, bronchial lavage water obtained by fibrobronchoscopy (bronchoalveolar lavage), as well as from (endo) tracheal, nasopharyngeal aspirate, sputum, biopsy or autopsy material of the lungs, whole blood, serum as well as in feces RT-PCR based methods are used. They are based on determination of RNA sequence encoding N gene, E gene, Orf1ab gene, RdRp gene or their parts. Among commercial kits, TaqMan 2019-nCoV Assay Kit v1 (Thermo Fisher Scientific, USA), Coronavirus (COVID-19) CE IVD (Primerdesign Ltd, UK), GSD NovaPrime® SARS-CoV-2 (COVID-19) (Gold Standard Diagnostics Corp, USA) and many others are available across the globe. Besides, countries are developing their own local solutions to provide permanent work for testing laboratories and to be independent of import processes (Labotaq SL, Spain; Biosan, Latvia; GeneProod, Czekh Republic). In Russia, commercial tests based on Orf1ab gene fragment detection have been officially registered and are used in laboratory routines (Kuzubov, 2020).

The objectives of this study were i) to simplify and to improve the WBE approach for SARS-CoV-2 gene copy number estimations to enable utilization in daily routines; and ii) to test this approach by monitoring the epidemiological situation in a typical large Russian city. For the first objective, the steps of sample concentrating and obtaining of qualitative information about viral particles were excluded from the analysis procedure. In addition, a locally produced and easily available commercial medical kit (Kuzubov, 2020) was used for RT and PCR reactions. We hypothesized that the approach of finding the lowest dilution that can be qualified as coronavirus positive, without sample concentrating would be more relevant and equally sensitive in terms of the determination of the COVID-19 epidemiological situation as compared to other WBE approaches used nowadays. For the second objective, samples were collected from 11 sewage inspection chambers in the city of Kazan which has 1.17 million citizens. The results obtained represent the first report about wastewater SARS-CoV-2 viral load in Russia.

## Materials & Methods

Two groups of samples were used in this study: i) feces and urine from ten COVID-19 patients from Kazan (Tatarstan, Russia) collected for 24 h, on the 5^th^-8^th^ day after registration of the first symptoms (Table 1); and ii) wastewater samples sampled in the sewage inspection chambers in Kazan (Tatarstan, Russia) on March 30^th^ and July 30^th^ 2020. The first group of samples was further used for the modelling experiment, and the second one was analyzed for SARS-CoV-2 particle presence.

### 2.1.1 Design of the modelling experiment

For the model experiment, feces and urine were collected separately from infected people during a 24 h period. After registration of mass and volume of excreta and sampling 1 g each for further RNA analysis, feces and urine were put into a 10 l plastic container with 5 l of model wastewater. The model wastewater contained (in 1 l): 0,05 g tooth paste, 0,25 g meat soup, 0,5 g of vegetable salad, 0,08 g of shower gel, 0,08 g of sand, 0,03 g of soil, 0,08 g of liquid soap, 0,08 g of dish soap and 0,03 g of washing powder. The excreta were thoroughly mixed with model wastewater using an electric drill with mixing bit at 1000 rpm. Furthermore, a 50 ml aliquot of the mixture was diluted 40 times to simulate the average daily volume of wastewater produced by a person living in a block of flats in Russia which is 200 l (model sample 1). Furthermore, serial dilution of the model sample was conducted to reach the final concentrations of model sample 1 from 25% to 0.005%. All the dilutions were prepared using model wastewater. Each sample and dilution was analyzed for the presence of SARS-CoV-2 particles using PCR based tests immediately after preparation. The analyses were conducted in triplicate.

### 2.2.2 Wastewater samples

Wastewater was sampled in Kazan on the 30^th^ of March and 30^th^ of July 2020 in 11 sewage inspection chambers, 10 of them were situated in the city’s residential areas and one in the city center (Figure 1).

**Figure 1.**
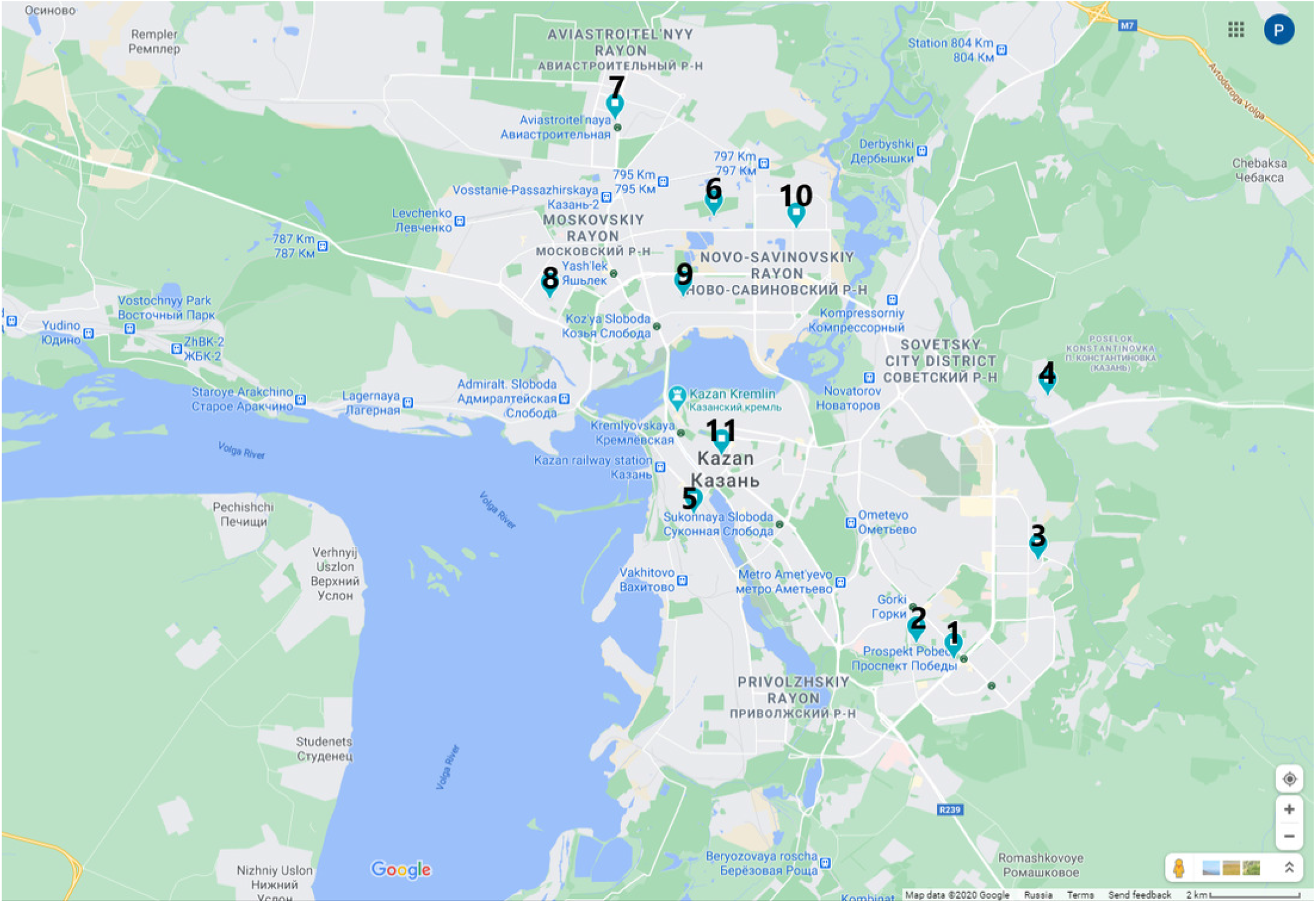
Location of wastewater sampling points in the city of Kazan (Russia)

Samples from each inspection chamber were collected during a 24 hour period, 200 ml each hour, mixed together in 5 l sterile plastic bottles. Between the collections, the bottles were stored at 4 °C. After finishing the sampling, the bottles were transported to the lab on ice and analyzed immediately. Each sample was analyzed in its initial state, as well as being diluted by 5, 25, 50, 75, 100, 150, 200 and 250 times with model wastewater if the initial sample was SARS-CoV-2 positive. Each initial and diluted sample was analyzed three times.

### 2.2. SARS-CoV detection

RNA was directly extracted from the samples using the viral RNA mini kit (QIAGEN, Germany). Detection of nucleic acid fragments of the coronavirus SARS-CoV-2 RNA was performed by the RT method combined with RT (RT-qPCR) using RNA SARS-CoV-2 by RT-qPCR Kit “OM-Screen-2019-nCoV-RT” (Syntol LLC, CEO, Russia). The amplification in this commercial kit uses two oligonucleotide primers flanking the Orf1ab gene fragment of the genome SARS-CoV-2. The kit contains 0.2 ml stripped PCR tubes with lyophilized reaction mixture. The kit allows simultaneous detection of the coronavirus SARS-CoV-2 RNA (R6G detection channel) and checking of the efficiency of nucleic acid extraction, the degree of inhibition of the reverse transcription and amplification reactions (FAM and Cy5 detection channels respectively). The RT-PCR reaction was conducted using the following temperature program on CFX-96 thermal cycler (Bio-Rad, USA): reverse transcription at 50°C for 15 min, inactivation at 95°C for 5 min followed by 50 three-step cycles of 95°C for 10 s, 58°C for 10 s, and 72°C for 20 s. According to the “OM-Screen-2019-nCoV-RT” Kit instructions, the wastewater samples were registered as positive when Ct in the R6G detection channel was less than 30. For feces and urine, the number of viral particles was calculated using an external control. For that, mouse hepatitis virus was used.

## Results

In this study, we aimed to develop a simplified approach for WBE of COVID-19 that can be applied in Russian cities. In order to do this, modelling with feces and urine of coronavirus infected people was conducted. 10 patients with differing ages and symptoms participated in the modelling experiment (Table 2). It was found that their urine did not contain virus particles while feces did. This observation is in line with results published by other researchers (Wölfel et al., 2020; Kitajima et al., 2020; Bhowmick et al., 2020). It should be noted that no dependence between viral RNA copy numbers and age of patients as well as duration and severity of their symptoms was found (Supplement Table S1).

In addition, feces and urine of each patient collected during 24 h were mixed with model wastewater, to simulate the dilution of excretions up to the final volume 200 l per day per person. This volume corresponds to average amount of waste water produced by residents of blocks of flats in central Russia (SNiP 2.04.01-85). The viral load in the resulting mixture theoretically should correspond to the wastewater from settlements with 100% infection rate. The resulting mixture was further diluted by model wastewater to simulate lower infection rates in the community. In both the mixture and in its solutions viral RNA was detected using RT-PCR method (Table 2).

Modelling presented in Table 2 demonstrates that the method of WBE used can detect a minimal rate of 10-2% of COVID-19 infection in the community. In contrast to the procedures of SARS-CoV-2 viral particles detection used by other authors, the modelling approach used in this study excludes the procedure of sample concentrating, the need and meaning of chosen external controls in RT-PCR (whole process control, molecular process control), and the recalculations of the viral particle numbers and of the infection rate (Wurtzer et al., 2020; Medema et al., 2020; Ahmed et al., 2020; Kitajima et al., 2020).

Using the results of modelling, SARS-CoV-2 viral load in the wastewater was analyzed in the city of Kazan, which is a typical million-citizen city in central Russia. Wastewater was sampled twice, at the beginning and at the peak of the epidemic, in ten sewage inspection chambers situated in residential areas as well as in one inspection chamber in the city center collecting wastewater from Kazan Federal University campus. The samples were characterized only qualitatively (coronavirus positive or negative), and if the SARS-CoV-2 particles were registered in the initial samples, they were diluted 5-200 times by model wastewater and reanalyzed (Supplement Table S2). The infection rate of people whose excreta are collected in the “positive” inspection chamber was then calculated from the lowest dilution found to be virus positive.

The results are presented in Table 3. As follows from the table, no SARS-CoV-2 particles were found in the Kazan wastewater in March while some were found in July. Four out of eleven initial wastewater samples taken in July were characterized as coronavirus positive. The lowest dilutions of these samples that were also found to be positive were 1:100, 1:150, 1:75 and 1:150 for samples 6, 7, 10 and 11, respectively, that corresponds to the 1.00%, 1.50%, 0.75% and 1.5% sickness rate in the residences that use the sewage inspection chamber. Taking into account the number of apartments in the blocks presented in Table 3 and the average number of people per apartment (3 people) (Draft master plan…, 2019) for the inspection chambers No 1-10, as well as the number of people that were present in the buildings using inspection chamber No 11, it can be calculated that the total number of people whose excreta were analyzed in the investigation was 1790, and the number of infected people was 8. That corresponds to 0.44% infection rate in the community.

Interestingly, the 11th sample was taken from the sewage inspection chamber that collects wastewater from 3 buildings of the Kazan Federal University campus in the center of Kazan. On the date of investigation, 65 people were present in the buildings, and all of them were interviewed for coronavirus symptoms and checked for body temperature. Despite the presence of viral particles in the wastewater, none of people reported feeling sick or having a fever. In the 5 days after this investigation, 1 case of COVID-19 was registered in one of these three Kazan Federal University buildings, and in the 7 days after investigation - 3 more new cases. The prognostic ability of WBE for COVID-19 investigations has been described by other authors. SARS-CoV-2 particles were found in the wastewater of Amersfoort in Netherlands on the 5th March 2020, while the first cases were registered only 6 days later (Medema et al., 2020).

## Discussion

In this study, the approach of serial dilution preparation for wastewater investigation for SARS-CoV-2 viral load was suggested. In the modelling experiment with the excreta of ten COVID-19 patients, it was demonstrated that the minimal rate of infected people in the community that can be detected by this method is 10-2%. This rate is in range of results presented in the literature.

Thus, it was demonstrated that the number of SARS-CoV-2 virus particles in wastewater ranges between 10^2^-10^4^ copies·l^-1^, and in feces from 10^5^-10^10^ copies·g^-1^ (Wu et al., 2020; Wurtzer et al., 2020; Wölfel et al., 2020; La Rosa et al., 2020; Farkas et al., 2020; Haramoto et al., 2020; Kitajima et al., 2020; Bhowmick et al., 2020). Taking into account the reported volume of wastewater produced by citizens (from 200 to 600 l), it can be calculated that the lowest detectable infection rate using WBE methods ranges between 10^−1^ and 10^−4^ % (Hata et al.; Wurtzer et al., 2020; Ahmed et al., 2020).

However, the classical WBE approach that includes concentration of the sample, PCR based detection and calculation of the viral particle numbers has several limitations. There are additional limitations especially for SARS-CoV-2 viral particle determination, since this virus is an RNA-based one, and therefore RT reaction is included in the determination procedure, and the probability of RT efficiency may also differ depending on many factors. Thus, it has been shown that the detection efficiency of SARS-CoV-2 viral particles ranges from 8.5 to 71.6% depending on the concentration method and the volume of the concentrated sample (Hata et al.; Medema et al., 2020; Haramoto et al., 2020; Kitajima et al., 2020). The results obtained may be dependent on molecular process control that can strongly vary in different lab protocols and influence the final results. As such, a control of other coronaviruses as well as mouse hepatitis virus, coliphage MS2 (ATCC 15597-B1), tobacco mosaic virus, Pseudomonas phage Φ6 and Murine norovirus have been used in different studies (Hata et al.; Medema et al., 2020; Haramoto et al., 2020; Kitajima et al., 2020). The complexity of the wastewater matrix also limits the detection accuracy SARS-CoV-2. For example, organic components like fat, protein or humic substances are reported to influence the efficiency of RT and PCR reaction. Besides, viruses of the same family or genus can interfere during RT-PCR reaction (Sims & Kasprzyk-Hordern, 2020; Farkas et al., 2020; Mao et al., 2020). It can be concluded that the results expressed in viral particle numbers found in the wastewater being compared with the viral particle numbers in feces may give incorrect information about the sickness rate in the community. Indeed, authors report controversial data about minimal number of COVID-19 infected people that can be detected using WBE – ranging from 1 to 100 cases per 100000 people (Wurtzer et al., 2020; Ahmed et al., 2020).

The “calibration table” obtained in the modeling experiment as well as the principle of samples’ serial dilution down to the lowest level where viral particles can be detected were used to investigate the wastewater produced in the city of Kazan situated in the European part of Russia. The residential districts of this city are typical for Russian cities of around one million population such as Nizhniy Novgorod, Novosibirsk, Volgograd and Samara as well as for larger cities Saint-Petersburg and Moscow. The procedure of SARS-CoV-2 particles’ detection in the wastewater included RNA extraction using viral RNA mini kit (QIAGEN, Germany), without the step of concentration, and RT-PCR reactions using widely available, locally produced kit for investigations of patient saliva and throat swabs (Kuzubov, 2020). These kits and simplifications were used in order to enable the easy inclusion of WBE of COVID-19 in the daily routine of wastewater plants and municipalities. The total duration of the detection procedure was about 2.5 h for 30-50 samples. Dilution of the samples was done using the model wastewater that was used to create the “calibration curve”, there was no need to use the external controls, which made the probability of obtaining the relevant results higher. Using the serial dilution approach, eleven wastewater samples were investigated twice, in March and July 2020. It was found that the level of viral particles in March was not detectable, while that in July was positive in four of eleven samples. This corresponds to the growth in numbers of registered COVID-19 cases in the city according to the official statistics (“Covid-19 official statistic,” 2020). After dilution of the positive samples, it was calculated that the average sickness rate of people connected to the inspection chambers under investigation was about 0.4%. This rate is much higher than that of officially registered cases which is 0.09% (“Covid-19 official statistic,” 2020). It could be explained by the statistical error due to the small number of inspection chambers investigated, by the high percentage of asymptomatic cases (Haramoto et al., 2020; Mao et al., 2020) especially in the Russian Federation (up to 70%) (Sharov, 2020) as well as by a special mentality. Usually, in Russia people report that they are ill only in cases of emergency, in other cases they prefer to stay home trying to take care of themselves. Further investigations that include more people taking part in a modeling experiment as well as more sampling of sewage inspection chambers are needed to obtain relevant information about the COVID-19 epidemiological situation in Russian cities using WBE. However, this study demonstrates the potential of the new approach based on serial dilutions for SARS-CoV-2 detection in wastewater.

## Conclusions

This is the first study that reports the detection of SARS-CoV-2 in wastewater in Russia using RT-PCR assay. The proposed approach to determining the presence of viral particles in wastewater makes it possible to detect more than 1 COVID-19 infected person per 10000 people with sufficient ease of analysis. The developed approach of detection of SARS-CoV-2 in wastewater due to dilution of model wastewater can be adapted for any types of wastewater collection, climatic conditions, commercial kits for DNA/RNA extraction and commercial kits for RT-PCR reaction.

## Data Availability

All data from the manuscript are available in case of reference to the authors, commercial use of the data from this scientific research is excluded

## Acknowledgements

Pavel Perfilov is acknowledged for inspiring this research.

